# RGnet: Recessive Genotype Network in a Large Mendelian Disease Cohort

**DOI:** 10.1101/2024.12.02.24318353

**Authors:** Fandi Ai, Lu Kang, Jiayi Zeng, Mingmin He, Mingjun Zhong, Jing Cheng, Yu Lu, Huijun Yuan, Fengxiao Bu

**Affiliations:** Institute of Rare Diseases, West China Hospital, Sichuan University, Chengdu 610041, China; BGI Research, Chongqing 401329, China

## Abstract

Recessive genotypes, including compound heterozygotes and homozygotes formed by rare variants that impact gene function, affect both alleles and were linked to numerous diseases and traits. However, the underlying patterns and interconnections of these recessive genotypes in large cohorts have rarely been studied. To address this gap, the Recessive Genotype Network (RGnet) was developed. This network model maps variant and genotype features to visualize and analyze recessive genotype patterns within large cohorts. Additionally, it uses permutation-based analyses to assess the enrichment of these genotypes in relation to specific phenotypes. Demonstrated through its application to the genetic deafness gene *SLC26A4* in 22,125 cases affected by hearing loss, RGnet successfully identified pathogenic variants with high connectivity, providing a reliable method for exploring the pathogenic mechanisms underlying recessive disorders or traits.

**Availability and Implementation:** RGnet is available from GitHub at https://github.com/jiayiiiZeng/RGnet

**Supplementary information:** Supplementary data are available at Bioinformatics online.

## 1 Introduction

Mendelian diseases are hereditary disorders caused by mutations or functional defects in a single gene, following predictable inheritance patterns—either autosomal or sex-linked, and dominant or recessive. Among these, recessive inheritance is particularly significant, accounting for approximately 50% of all genetic disorders and phenotypes, as reported by the Online Mendelian Inheritance in Man (OMIM) database^1^. A defining feature of recessive genetic disorders is that affected individuals must carry pathogenic variants in both alleles of the same gene. These recessive genotypes (RG) are further categorized into homozygotes, in which individuals possess two identical mutated alleles, and compound heterozygotes, where individuals have different mutations on each allele of the disease-causing gene. Understanding the precise nature of these variants is crucial for accurately identifying pathogenic mechanisms in Mendelian diseases.

In most cases, recessive genetic disorders present sporadically, affecting isolated individuals or siblings within a nuclear family, with large families exhibiting multiple affected members being rare, except in populations with high rates of consanguinity. Due to the complex nature of recessive inheritance, determining the pathogenicity of recessive genotypes often necessitates data beyond a single family’s segregation patterns. Instead, it requires evidence from multiple, unrelated affected families to strengthen the conclusions about a variant’s pathogenicity. The ACMG/AMP guidelines for variant classification, specifically the PM3 criterion, reflect this need by incorporating evidence based on the presence of RGs in numerous unrelated cases ^2^. This approach helps to establish a continuum of pathogenicity evidence, ranging from supporting to very strong, depending on the consistency and frequency of the observed genotype-phenotype correlations. Larger, disease-specific patient cohorts have been shown to significantly enhance the accuracy and reliability of RG-based pathogenicity assessments, emphasizing the importance of broad datasets in genetic research^3,4^. However, the ability to accurately classify variants as pathogenic, particularly in the context of recessive inheritance, remains a complex and evolving area of study.

In recent years, advancements in high-throughput sequencing technologies have revolutionized the study of genetic disorders, leading to the emergence of large cohort studies focused on rare diseases. Notable examples include the Deciphering Developmental Disorders (DDD) project ^5^, the UK-100K initiative ^6^, and the Chinese Deafness Genetics Consortium (CDGC)^7^. These studies have provided unprecedented insights into the genetic basis of rare disorders by enabling the comprehensive analysis of large patient cohorts. Additionally, multicenter initiatives such as the Centers for Mendelian Genomics (CMG) ^8^and the Finding of Rare Disease Genes (FORGE) ^9^ project have established collaborations for rare disease genetic testing, facilitating the aggregation of sequencing data and diagnostic information from multiple institutions. These efforts have significantly contributed to the interpretation of pathogenic variants and the identification of novel disease-causing genes. Despite these advances, there remains a critical gap in the tools available for analyzing compound heterozygous and homozygous variants within large genetic datasets. Existing tools, such as those based on mutation-rate-driven RG count calculations^3,10^, BeviMed^11^, and SAIGE-Gene+ ^12^, are primarily designed for gene association studies in recessive traits. While these tools have been instrumental in identifying gene-disease associations, they are often limited in their ability to effectively analyze the complex patterns of compound heterozygosity and homozygosity that characterize many recessive disorders, particularly in large, diverse cohorts. Given the growing availability of large, disease-specific patient cohorts, there is an urgent need for new analytical tools that can more effectively discover RG distributions and interconnections, refine the variant pathogenicity interpretation, and support the identification of novel disease-causing genes. Accurate and efficient analysis of RGs in these cohorts is essential for advancing our understanding of the genetic basis of monogenic diseases and improving the accuracy of genetic diagnoses.

To address these challenges, this study introduces RGnet, a novel tool designed to illustrate RG relationships within patient cohorts and calculate RG enrichment at both the variant and gene levels. RGnet provides researchers with a precise method for identifying pathogenic variants in recessive inheritance patterns by integrating data from large cohorts. By enhancing the ability to analyze RGs, RGnet aims to advance the study of monogenic diseases, facilitate the identification of novel pathogenic variants, and ultimately contribute to the development of more effective diagnostic and therapeutic strategies.

## 2 Materials and methods

The RGnet model consists of four primary modules: variant preprocessing, variant phasing, network construction, and RG enrichment analysis. Each RGnet represents the RG patterns of a recessive gene within a cohort (Figure 1A). In the network, each variant is represented as a node; a compound heterozygote in an individual is denoted by an edge connecting two nodes, while a homozygote is depicted as a self-looping node. RGnet translates the biological properties of the RGs in the cohort into network features, such as the total number of variants (number of nodes), the overall count of RGs (total node weight), RG count per variant (node weight), and RG frequency (edge weight). Additionally, RGnet uses a permutation approach to estimate the expected RG count, assuming RG occurrences result from random mating in a large, unrelated population without selection, genetic drift, or gene flow. By comparing observed and expected RG counts, RG enrichment analysis evaluates the association between phenotypes and specific variants or genes.

**Figure 1.**
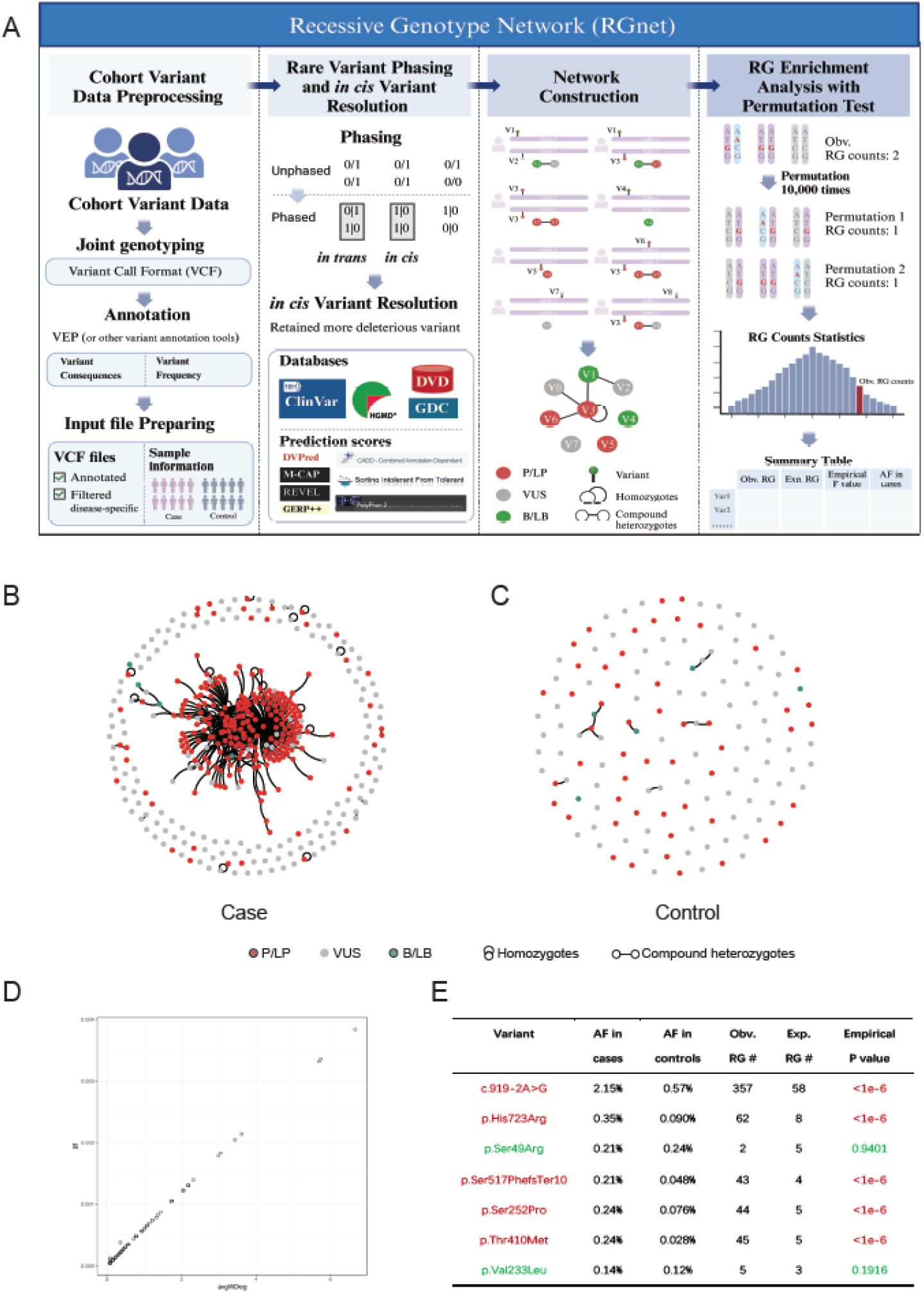
RGnet Construction and Enrichment Analysis. **A:** Scheme of RGnet construction; **B:** Network of *SLC26A4* for the cases selected from CDGC. Nodes represent individual variants, with edges indicating connections between variant pairs. Nodes are color-coded: red for Pathogenic/Likely Pathogenic (P/LP), gray for Variants of Uncertain Significance (VUS), and green for Benign/Likely Benign (B/LB); **C:** Network of *SLC26A4* for the controls selected from CDGC; **D:** The correlation between the average weighted degree of node and variant allele frequency (AF); **E:** Enrichment analysis of selected pathogenic and benign variants in the *SLC26A4* gene showing the correlation between elevated RG count and variant pathogenicity.

### 2.1 Cohort Variant Data Preprocessing

RGnet takes a cohort Variant Call Format (VCF) file as input. Variant annotation is performed using VEP or similar tools. To analyze phenotypes associated with recessive genotypes, it is essential to apply filtering to retain relevant variants. For example, in the analysis of genetic hearing loss, variants with a minor allele frequency (MAF) of less than 0.005 and significant functional impacts, such as protein-truncating variants, missense mutations, and splicing changes, are retained. Other information, including sample data, gene details, and genotype information, was collected and processed for the subsequent steps.

### 2.2 Rare Variant Phasing and *in cis* Variant Resolution

To minimize the impact of *in cis* variants on the analysis of compound heterozygotes, variant phasing is essential. Physical, family-based, and population-based phasing methods can be utilized for this purpose. In this study, combination of trio-based phasing, read-based phasing and an expectation-maximization phasing algorithm were integrated to analyze variant pairs, inferring the phase of each pair within the same gene from the input VCF file ^13^. For each pair of *in cis* variants, the variant with the stronger pathogenicity or more severe functional impact was retained. This determination was based on variant databases such as ClinVar or the Human Gene Mutation Database (HGMD), or on in silico prediction scores (e.g., REVEL). If both variants exhibited identical pathogenicity, one was randomly excluded. Finally, only compound heterozygotes and homozygotes were retained for further analysis.

### 2.3 Network Construction

The VCF file was partitioned by gene, and each subset was converted into a connectivity matrix to build the RGnet, allowing for a clear visualization of RG network patterns within the cohort. In the network, edges represent compound heterozygous relationships between two variants, while self-loops denote homozygous variants. The weight of each edge corresponds to the RG count, reflecting how often a pair of variants forms an RG within the cohort. Similarly, node weights represent the variant allele counts, indicating the frequency of each variant. To further enhance the interpretability of the network, nodes representing pathogenic (P) and likely pathogenic (LP) variants, as defined by ClinVar or HGMD, are visually distinguished using specific colors. This color-coding allows researchers to quickly identify variants with higher clinical significance within the network. The resulting RGnet provides a comprehensive overview of the distribution and relationships of RGs, enabling deeper insights into recessive inheritance patterns and potential genotype-phenotype correlations.

### 2.4 RG Enrichment Analysis with Permutation Test

A permutation test was conducted to estimate the expected RG count for each variant in the cohort. First, the observed occurrence of RGs and individual variant frequencies were recorded. Then, 100,000 permutations were performed by randomly shuffling all alleles (2N) across all samples (N), ensuring that the occurrence of each variant remained consistent with the original data, thereby preserving the characteristics of the variant dataset. For each permutation, the RGnet was reconstructed, and the number of compound heterozygotes and homozygotes was recorded. After the permutations, the distribution of RG counts for each variant was analyzed. The average RG count across all permutations represented the expected RG count. To assess statistical significance, the empirical p-value (percentile rank) of the observed RG count was calculated within the permutation distribution. The final output included a network graph and a comprehensive summary of key results, including variant population frequencies, observed RG counts, mean RG counts from the permutations, and the corresponding p-values. This output provides a rigorous framework for detecting statistically significant RG enrichment, thereby supporting the interpretation of recessive inheritance patterns within the cohort.

## 3 Application and Results

RGnet was applied to the *SLC26A4* gene using data from the CDGC cohort. Cases were included based on the following criteria: (1) underwent high-throughput sequencing involving the *SLC26A4* gene; (2) exhibited a recessive inheritance pattern or had no recorded family history of hearing loss; and (3) were not genetically diagnosed with dominant or X-linked deafness genes. This selection yielded 12,304 hearing loss cases and 7,524 controls. Variants were retained if they met the following criteria: (1) passed quality checks with genotype quality > 20 and depth > 10; (2) had a minor allele frequency (MAF) < 0.5% in both CDGC controls and gnomAD populations; and (3) were predicted to affect protein function (e.g., missense, frameshift, stop-gain, inframe deletions/insertions, start-loss, stop-loss) or splicing (within 1-3 bases of an exon or within ±20 base pairs of an intron). Additionally, all pathogenic/likely pathogenic (P/LP) variants curated by the CDGC workgroup were retained. After phasing and collapsing *in cis* variants, 498 variants were included for RGnet analysis.

Network construction and enrichment analysis were performed separately on the case and control datasets, resulting in two distinct network graphs (Figures 1B and 1C). The results revealed significant structural differences between the networks for the *SLC26A4* gene in the cases and controls. In the case network, 365 types of RGs were identified with a weight of ≥1, including 84 types with a weight of ≥2. In contrast, the control network contained only 11 types of RGs, all with a weight of 1. The case network also featured several highly connected central variants (nodes), distinguishing it from the control network.

Most P/LP variants in the case network exhibited higher observed RG counts than expected, with significant p-values (Figure 1B and 1E). For instance, the variant c.919-2A>G had the highest observed RG count of 357, far exceeding the expected RG count of 58, underscoring its pathogenic role (Figure 1E). In contrast, benign or likely benign (B/LB) variants and variants of uncertain significance (VUS) showed observed RG counts that closely aligned with the expected values (Figures 1B and 1E). These differences highlight a clear correlation between RG enrichment and variant pathogenicity.

Additionally, we observed a linear correlation between expected RG counts (mean weighted degree of nodes) and variant allele frequency (Figure 1D). As variant allele frequency increased, the number of observed RGs correspondingly rose. Therefore, when interpreting variant pathogenicity based on observed RG counts, as suggested by the PM3 rule, allele frequency should be taken into account.

## 4 Conclusion

RGnet demonstrated to be a valuable tool for visualizing RG patterns in large patient cohorts. It facilitates the interpretation of variant pathogenicity, both by aligning with ACMG/AMP guidelines, which involve counting neighboring nodes with varying levels of pathogenicity, and through permutation tests that assess RG enrichment. The application of RGnet to the *SLC26A4* gene highlights its effectiveness in identifying pathogenic variants. Ultimately, RGnet provides researchers with a precise and reliable method for investigating the pathogenic mechanisms of rare diseases under recessive inheritance, with significant potential for broader applications. RGnet is available on GitHub at [https://github.com/jiayiiiZeng/RGnet].

## Data Availability

All data produced in the present study are available upon reasonable request to the authors

## Acknowledgements

We would like to extend a special thanks to Information Center of West China Hospital for technical support.

## Data availability

The ClinVar data was downloaded from https://ftp.ncbi.nlm.nih.gov/pub/clinvar/vcf_GRCh38/. The HGMD dataset can be accessed at https://www.hgmd.cf.ac.uk/ac/index.php.

